# Previous visceral leishmaniasis relapses outperform immunological or clinical signs as predictors of further relapses in patients co-infected with HIV-1

**DOI:** 10.1101/2021.12.14.21267767

**Authors:** Yegnasew Takele, Tadele Mulaw, Emebet Adem, Rebecca Womersley, Myrsini Kaforou, Susanne Ursula Franssen, Michael Levin, Graham Philip Taylor, Ingrid Müller, James Anthony Cotton, Pascale Kropf

## Abstract

Visceral leishmaniasis (VL) and HIV co-infection (VL/HIV) has emerged as a significant public health problem in Ethiopia, with up to 30% of VL patients co-infected with HIV. These patients suffer from recurrent VL relapses and increased mortality. Our aim was to assess whether VL/HIV co-infected patients with a previous history of VL relapses (recurrent VL/HIV) have a poorer prognosis as compared to HIV patients presenting with their first episode of VL (primary VL/HIV). Our results show that recurrent VL/HIV patients have a higher parasite load, higher mortality and that their relapse-free survival is significantly shorter. The poorer prognosis of recurrent VL/HIV patients is accompanied by lower weight gain and lower recovery of all blood cell lineages, as well as lower production of IFNγ, lower CD4^+^ T cell counts and higher expression levels of PD1 on T cells. Furthermore, our results show that prior history of VL relapse is an important risk factor for future relapse. Both CD4^+^ T cell count and parasite load were also associated with a higher risk of VL relapse, but neither of these was independently associated with relapse risk after adjustment for previous VL relapse history.

We propose that in addition to the current treatments, novel interventions should be considered at the time of VL diagnosis in VL/HIV patients; and suggest that improved anti-leishmanial and antiretroviral treatments, as well as immune therapy, through PD1/PDL-1 blockade and/or through IFNγ administration, could result in more efficient parasite killing and thereby reduce the relapse rate and improve survival.

## INTRODUCTION

Visceral leishmaniasis (VL), also named kala-azar, is a potentially fatal neglected tropical disease, caused by parasites of the genus *Leishmania*. An estimated 50,000 to 90,000 new cases of VL occur worldwide annually, but only 17,082 new cases of VL were reported in 2018, with Brazil, Ethiopia, India, South Sudan and Sudan, each reported >1000 VL cases, together representing 83% of all cases globally (1). Because of the remote location of VL endemic areas and the lack of surveillance, it is widely accepted that this is a vast underestimation of the real burden of VL in endemic areas. VL imposes a huge pressure on the developing countries and delays economic growth, with an approximate annual loss of 2.3 million disability-adjusted life years (2).

In Ethiopia, where this study took place, VL is one of the most significant vector-borne diseases: over 3.2 million people are at risk of infection (3). VL is caused by infections with parasites of the *Leishmania* (*L.*) *donovani* species complex. Not all infected individuals will develop the disease: some will stay asymptomatic, but in those who develop VL, the disease is characterised by hepatosplenomegaly, fever, pancytopenia and severe weight loss; this stage of the disease is mostly fatal if left untreated (4–6). Following the HIV-1 pandemic, VL has emerged as an opportunistic infection: HIV infection increases the risk of developing symptomatic VL and VL accelerates the progression of HIV infection to AIDS (7, 8). HIV co-infection presents a significant challenge in the prevention and control of VL (9, 10): VL/HIV co-infected patients experience increased rates of VL relapse, mortality and treatment failure compared to patients with VL alone (10–12). Our knowledge of the immunology of VL/HIV co-infections is still limited. Increased levels of soluble CD40L, a molecule previously associated with the resolution of VL (13) and lower levels of neopterin, a molecule associated with activation of macrophages, were present in the plasma of HIV/VL as compared to HIV patients (14). Van den Bergh *et al.* suggested that myeloid derived suppressor cells might play a role in severely immunocompromised HIV/VL co-infected patients (15). We have shown previously that the activity of arginase, an enzyme with immunoregulatory properties, was significantly higher in the blood of VL/HIV co-infected patients as compared to VL patients, indicating that increased arginase activity contributes to T cell hyporesponsiveness in VL/HIV co-infected patients (16). A recent study has identified a 4-gene immune signature that could identify patients who fail treatment from those who are successfully treated (17). Santos-Oliveira *et al.* showed that increased numbers of activated T lymphocytes are present in VL/HIV co-infected patients, even in those with undetectable viral load (18). This increased in T cell activation was associated with LPS levels (19). Low CD4^+^ T cell counts are also a hallmark of VL/HIV patients (5, 20, 21).

We have recently shown that throughout follow-up, as compared to VL patients, VL/HIV patients display (12):

- higher parasite load;
- impaired antigen-specific IFNγ production by whole blood cells;
- lower CD4^+^ T cell counts;
- higher PD1 expression on CD4^+^ and CD8^+^ T cells.

Our data also show that 78.1% of VL/HIV patients experience at least one relapse (12). Reliable data on both clinical and immunological parameters from VL/HIV patients who experience VL relapse are still limited: the following markers have been shown to predict VL relapse in VL/HIV patients:

- failure to clear parasite load after anti-leishmanial treatment (20), as well as presence of circulating *Leishmania* DNA (22);
- low CD4^+^ T cell counts (5, 20);
- previous VL episodes in VL/HIV patients (5, 20, 23), though a study showed no association between a previous history of relapse and increased risks of future relapse (24).

In a study where VL/HIV patients received VL prophylaxis treatment after initial cure of VL, it was shown that the group of patients who did not relapse displayed lower soluble CD14 and anti-*Leishmania* IgG3 levels, as well as less activated T cells, suggesting that these patients could control immune activation more efficiently (25). Recently a study assessed the involvement of the thymus in the replenishment of T cells: their results suggest that in VL/HIV co-infected patients who do not relapse, more new emigrant T cells can be detected that might contribute to the control of parasite replication (26). The work by Casado *et al.* also showed that VL/HIV patients had an even worse immunological status as compared to ART treated HIV patients who failed to improve their CD4^+^ T cell counts (27). In our recent study, we showed that VL/HIV patients who did not relapse after initial clinical cure had lower parasite loads as measured by RNAseq in whole blood, produced higher levels of antigen-specific IFNγ, maintained higher CD4^+^ and CD8^+^ T cell counts and lower PD1 expression on CD4^+^ and CD8^+^ T cells, as compared to VL/HIV patients who did relapse (12). In this study (12), out of the 49 HIV patients who presented with VL at the treatment centre, 21 (43%) patients presented with their first episode of VL (Primary VL/HIV patients) and 28 (57%) had a previous history of VL (recurrent VL/HIV patients). In the current study, we assessed the impact of a previous history of VL relapse on relapse-free survival, clinical parameters and the immune response and compared those to HIV patients presenting with their first episode of VL. To test this, we performed an extensive follow-up study of primary and recurrent VL/HIV patients, from the time of VL diagnosis to 6-12m post the end of treatment; determined the frequency of VL/HIV patients who remain relapse free over time, and identified clinical and immunological markers associated with VL relapse in these two cohorts.

## METHODS

### Patient recruitment

For this cross-sectional study, we followed the cohort of 49 VL/HIV patients (median age 33.5±1.0 years) that was described in (12). Twenty-one patients presented with their first episode of VL (primary VL/HIV, P VL/HIV) and 28 with at least one previous episode of VL (recurrent VL/HIV, R VL/HIV, Table 1). The diagnosis of VL was based on positive serology (rK39) and the presence of *Leishmania* amastigotes in spleen or bone marrow aspirates (28). The diagnosis of HIV was done in accordance with the Ethiopian National HIV Screening Test Guidelines (29). Forty-six VL/HIV patients were on anti-retroviral therapy (ART) at the time of VL diagnosis; the remaining three started ART at the end of the anti-leishmanial treatment. All treatments were administered according to the Guideline for Diagnosis, Treatment and Prevention of Leishmaniasis in Ethiopia (30) (Table 2). At the end of treatment, patients were discharged if they look improved, afebrile, had a smaller spleen size and an improved haematological profile (initial clinical cure). When there was no or insufficient clinical improvement, a tissue aspirate was performed to test for the presence of *Leishmania* amastigotes (Test Of Cure, TOC). If the TOC was still positive (i.e. incomplete cure), treatment was continued until TOC becomes negative or the patients have sufficient clinical improvement (30).

**Table 1:** recurrent VL/HIV patients: number of relapses at ToD.

| **Number of relapses** | **Number of patients** |
| --- | --- |
| 1 | 4 |
| 2 | 7 |
| 3 | 5 |
| 4 | 2 |
| 5 | 1 |
| 6 | 1 |
| 7 | 1 |
| 9 | 2 |
| 10 | 2 |
| 12 | 1 |
| 14 | 2 |

Number of previous relapses in R VL/HIV patients (n=28) at ToD when this study was started. ToD=time of diagnosis

**Table 2:** Type and duration of anti-leishmanial treatments.

| **Number of**<br>**P VL/HIV patients** | **Treatment** | **Duration of**<br>**treatment (days)** |
| --- | --- | --- |
| 1 | SSG+PM | 61 |
| 3 | SSG | 28 |
| 1 | AmBisome | 65 |
| 4 | AmBisome | 28 |
| 7 | AmBisome + Miltefosine | 28 |
| 1 | AmBisome + Miltefosine | 30 |
| 1 | AmBisome + Miltefosine | 40 |
| 1 | AmBisome + Miltefosine | 45 |
| 1 | AmBisome + Miltefosine | 62 |
| 1 | AmBisome + pentamidine | 30 |
| **Number of**<br>**R VL/HIV patients** | **Treatments** | **Duration of**<br>**treatment (days)** |
| 1 | SSG+PM | 61 |
| 1 | SSG | 28 |
| 1 | SSG | 71 |
| 2 | AmBisome | 28 |
| 1 | AmBisome | 65 |
| 10 | AmBisome + Miltefosine | 28 |
| 1 | AmBisome + Miltefosine | 45 |
| 1 | AmBisome + Miltefosine | 62 |
| 1 | AmBisome + Miltefosine | 90 |
| 1 | AmBisome + Miltefosine | 52 |
| 1 | AmBisome + Miltefosine | 101 |
| 1 | AmBisome + pentamidine | 30 |
| 1 | AmBisome + pentamidine | 62 |
| 5 |  | Treatment not<br>completed |

SSG= Sodium stibogluconate, PM= Paramomycin and AmBisome= Liposomal amphotericin

The definitions of no relapse and relapse are defined as follows:

- no relapse: absence of clinical features of the disease 6 months after completion of the recommended dose and duration for VL treatment.
- Relapse: patient with VL treatment history presenting with clinical visceral leishmaniasis symptoms and is diagnosed with positive parasitology after successful completion of the treatment.

Antiretroviral therapy (ART) was provided according to the National Guidelines for Comprehensive HIV Prevention, Care and Treatment (29, Table 3).

**Table 3:** ART.

| **Number of P**<br>**VL/HIV** | **ART at ToD** | **ART after EoT** |
| --- | --- | --- |
| 18 | TDF, 3TC and EFV |  |
| 1 | AZT, 3TC and EFV |  |
| 2 | Not on ART | TDF, 3TC and EFV |
| **Number of R**<br>**VL/HIV** |  |  |
| 25 | TDF, 3TC and EFV |  |
| 2 | TDF 3TC and ATV |  |
| 1 | Not on ART | TDF, 3TC and EFV |

TDF: Tenofovir disoproxil fumarate, 3TC: Lamivudine, EFV: Efavirenz, AZT:
azidothymidine, ATV: atazanavir. ToD=Time of Diagnosis; EoT=End of Treatment.

Patients were recruited at four different time points (12): time of diagnosis (ToD); end of treatment; 3 months after the end of treatment (3m); and 6-12 months after the end of treatment (6-12m). At the end of this 3-year study, all VL/HIV patients who had not relapsed during the 12 months follow-up were contacted by phone to find out if they had had any subsequent relapse.

### Sample collection and processing

Venous blood (13ml) samples were taken and distributed as follows:

- 2.5ml in PAXgene tubes for RNA sequencing
- 8 ml in heparinised tubes: 3ml for the whole blood assay (WBA), 5ml to purify PBMC (31) (flowcytometry) and plasma (cytokines)
- 2.5ml in EDTA tubes (whole blood for: CD4^+^ T cell count, white and red blood cell and platelet counts and plasma for viral load)

- WBA: soluble leishmania antigen (SLA) was prepared as described in (31).
- Flow cytometry: the following antibodies were used: CD4^FITC^, CD8^PE CY7^ and PD1^PE^ (eBioscience). The data on PD1 expression are shown as the integrated Median Fluorescent Intensity (iMFI: % PD1^+^ T cells x PD1 MFI) (32).
- CD4^+^ and CD8^+^ T cell counts: 100µl of blood was stained with CD4^FITC^, CD8^PE CY7^ and CD3^PerCP-eFluor® 710^ (eBioscience) for 15min at 4°C; red blood cells were lysed using BD FACS™ Lysing Solution for 5min at room temperature. Acquisition was performed using BD Accuri™ C6 flow cytometry and data were analysed using BD Accuri C6 analysis software.
- IFNγ and IL-10 levels were measured in the supernatant of the WBA using IFN gamma and IL-10 Human ELISA Kit (Invitrogen) according to the manufacturer’s instructions. The optical densities obtained with the unstimulated whole blood cells were subtracted from the optical densities obtained with whole blood cells stimulated with phytohaemagglutinin (PHA) or soluble *Leishmania* antigen (SLA).
- HIV viral load: plasma was isolated by centrifuging 2ml of blood and frozen at -80°C. HIV viral load was measured in the Central Laboratory of the Amhara Public Health Institute, Bahir Dar, by using Abbott RealTime HIV-1 Qualitative (m2000sp), according to the manufacturer’s instructions.
- White and red blood cell, and platelet counts were measured using a Sysmex XP-300T^M^ automated haematology analyser, (USA) following the manufacturer’s instruction.

### Relapse rate

Log-rank tests for duration of follow-up at event end points provided two-sided *p*-values; Kaplan-Meier curves are presented for visual interpretation. The primary outcome survival until three years of follow-up was completed; relapse was the only censoring event. Censoring events were reported at the pre-planned follow-up period (3, 6-12 and after 3 years) at which they were identified. Cox proportional-hazards regression analysis was used to estimate hazard ratios and 95% confidence intervals.s

### mRNA

RNA was extracted from 2.5 ml whole blood using the PAXgene 96 blood RNA kit (Qiagen) and globin mRNA was depleted using the GLOBINclear kit (Ambion). Sequencing libraries were prepared using the KAPA Stranded mRNA-Seq Kit (Roche) with 10 PCR cycles, then sequenced as 75bp paired-end reads on the Illumina HiSeq 4000 platform. Sequencing reads were mapped with Salmon v.1.30 against concatenated sequence of human gencode transcriptome release 34, transcripts for *L. donovani* LV9 from TriTrypDB release 46. Pseudo-counts were imported into R v4.0.3 using the tximport v1.18.0 and transformed into lengthscaledTPM. Total *Leishmania* expression was quantified as the sum across all LV9 transcripts with the exception of feature LdLV9.27.2.206410, an 18S rRNA gene to which human transcripts also map.

### Statistical analysis

Statistical tests are specified in the legend of each figure. The following were used: Mann-Whitney, Kruskal-Wallis and Fisher’s exact tests. Differences were considered statistically significant at *p*<0.05. *=p<0.05, **=p<0.01, ***=p<0.001 and ****=p<0.0001. Unless otherwise specified, results are expressed as median±SEM.

### Survival analysis

One-year relapse rates were estimated using the Kaplan-Meier method, with quantitative factors split into two categories at the median value, with non-VL death and loss to follow-up as censoring events at EoT, 3-, 6- or 12-months follow-up. To formally test the significance of differences in relapse we used the Cox proportional-hazards model, initially estimating the hazard ratio with single-factor models for each covariate, where covariates were considered significantly associated with relapse if the 95% confidence interval for the hazard ratio did no overlap 1. We attempted to build a multivariable Cox model for the three significant factors, but this was not possible due to the level of missing data for end-of-treatment CD4 cell counts, so we instead built two-factor models for VL history and the two other significant factors (parasite load at time of diagnosis and CD4 count at end-of-treatment) to estimate the adjusted hazard ratio for each factor, testing their significance using ANOVA. These survival analyses used v3.2.7 of the survival package (33) in R v4.0.2 (R Foundation for Statistical Computing, 2021 #3868)

### Study approval

This study was approved by the Institutional Review Board of the University of Gondar (IRB, reference O/V/P/RCS/05/1572/2017), the National Research Ethics Review Committee (NRERC, reference 310/130/2018) and Imperial College Research Ethics Committee (ICREC 17SM480). Informed written consent was obtained from each patient and control.

## RESULTS

### CLINICAL DATA

#### Frequency of VL relapse in VL/HIV patients with primary VL and multiple relapses

We previously reported that despite initial clinical cure, 78.1% of VL/HIV patients relapsed at least once (12). In this cohort of VL/HIV patients, 21 HIV patients presented with their first episode of VL (primary VL/HIV patients, P VL/HIV) and 28 presented with VL but had already experienced previous VL (recurrent VL/HIV, R VL/HIV) (Table 1, Figure 1). To assess if the absence of a previous history of VL results in a longer relapse-free survival, a better prognosis and a stronger immune response, these two cohorts of patients were followed for a period of 6-12 months (Figure 1) and detailed clinical and immunological data were collected. As shown in Figure 2, the percentage of relapse-free survival over a period of 12 months was significantly higher in P as compared to R VL/HIV patients (76.5% vs 21.1%, respectively, *p*=0.0011). R VL/HIV relapsed earlier as compared to P VL/HIV patients: 42.1% vs 11.8% at 3m, respectively.

**Figure 1:**
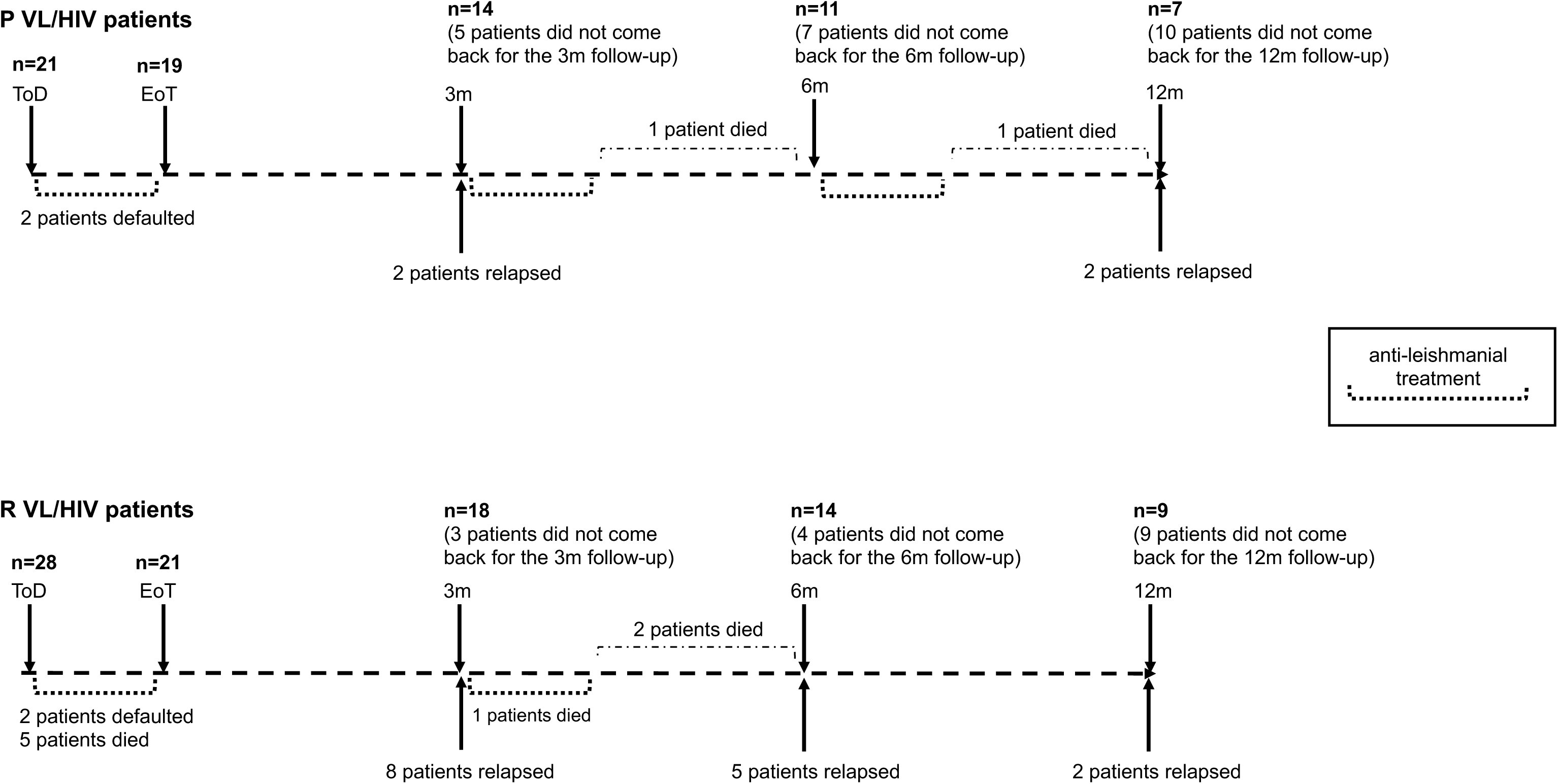
Follow-up of primary and recurrent VL/HIV patients: 21 P and 29 R VL/HIV patients were followed for up to 6-12m. ToD=Time of Diagnosis; EoT=End of Treatment; 3m=3 months post EoT; 6-12m=6-12 months post EoT.

**Figure 2:**
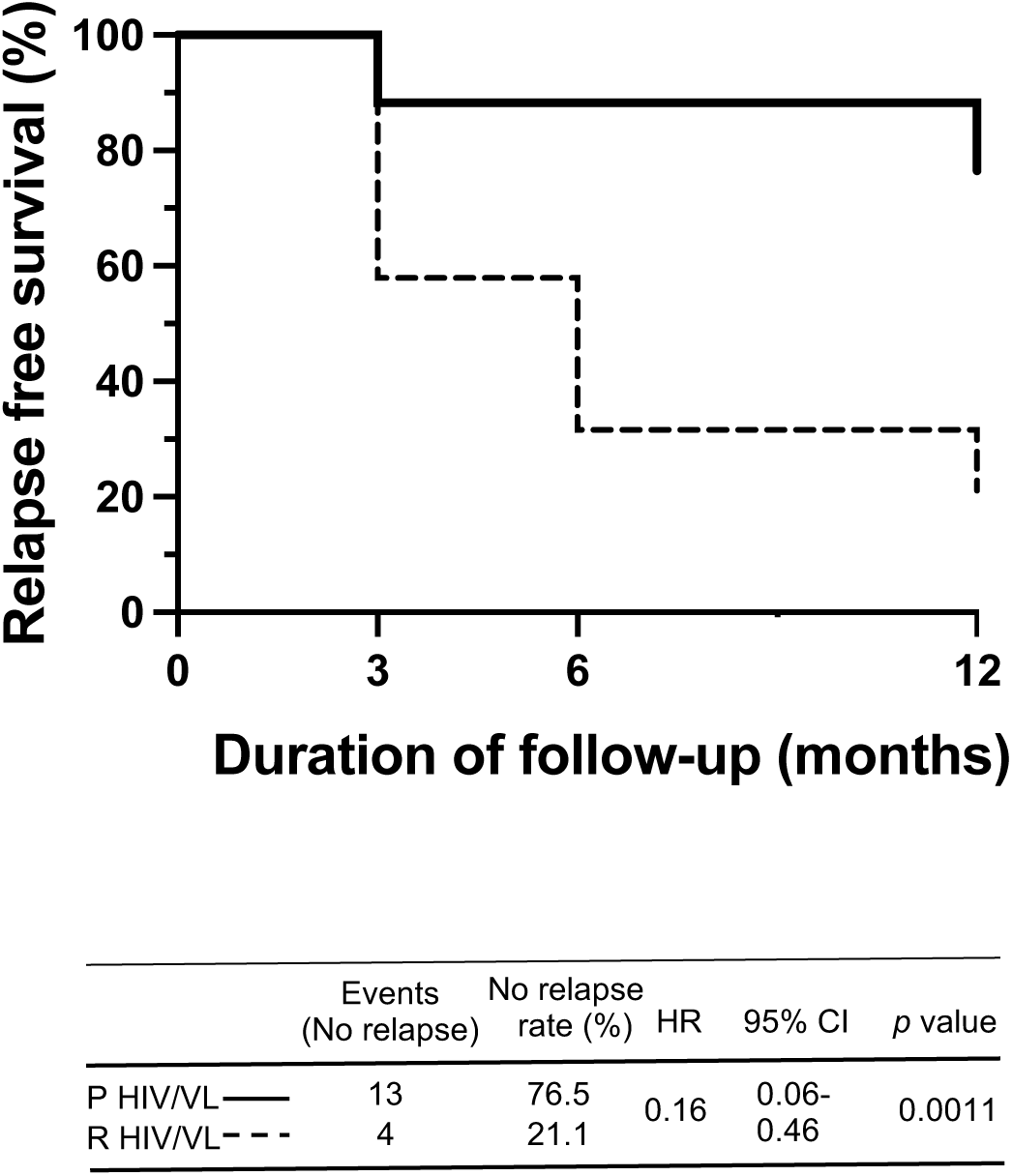
Relapse free survival: Kaplan-Meier curves of participant VL relapses comparing P VL/HIV to R VL/HIV patients. The hazard ratios (with 95% confidence intervals and *p* values) obtained from the Cox model indicated the change in relapse-free survival following treatment for VL for these groups. HR, hazard ration; CI, confidence interval.

Whereas we didn’t collect any clinical or immunological data after the initial 12-month follow-up, all VL/HIV patients who had not relapsed were contacted. A further four P VL/HIV and two R VL/HIV patients had relapsed (percentage of relapse-free survival: 23% and 15%, respectively, *p*=0.0327).

#### Parasite grades and viral load

Next, we compared parasite grades in splenic aspirates of P and R VL/HIV patients at ToD. As shown in Figure 3A, parasite grades were significantly lower in P than R VL/HIV patients (*p*<0.0001). Parasite grade can only be measured when the spleen is palpable and >3cm below the costal margin; it is therefore mainly measured at ToD. We have previously shown that RNAseq can be used to measure the total expression of *L. donovani* mRNAs (*Ld* mRNA) in blood (12). The median *Ld* mRNA was lower at ToD in P VL/HIV patients, this was not statistically significant (Table 4, Figure 3B). At the end of treatment and at 3m, there was significantly less *Ld* mRNA in P VL/HIV (Figure 3B, Table 4). At 6-12m, despite an increased median *Ld* mRNA in R HIV/VL patients, the differences between the 2 groups were not significant (Table 4, Figure 3B). As shown in Figure 2, significantly more P VL/HIV patients remain relapse-free during follow-up (3m and 6-12m time points). To assess whether these patients have a lower parasite load, we further divided each cohort of P and R VL/HIV patients into 2 subgroups: those who didn’t relapse and those who did relapse after initial cure. The *Ld* mRNA medians were lower in P and R VL/HIV who did not relapse during follow-up as compared to those who relapsed (Figure 3C). As expected, an increase in parasite load was observed in R VL/HIV patients who relapsed (Figure 3C). However, as shown in Figure 1, only four P VL/HIV patients relapsed during follow-up, however not enough blood could be collected from these four for all tests performed in our study and in this instance, PAXgene tubes were only collected from 2 patients; furthermore, only four R VL/HIV did not relapse, and PAXgene tubes were collected from three of them. It was therefore not possible to draw meaningful conclusions from these data.

**Table 4A:** *L. donovani* mRNAs.

|  | **P VL/HIV** | **R VL/HIV** | ***p* values** |
| --- | --- | --- | --- |
| **ToD** | 136±608 | 1786±1229 | 0.1068 |
| **EoT** | 0.1±0.4 | 1.5±341.4 | 0.0007 |
| **3m** | 0.1±149.3 | 1047±1188 | 0.0099 |
| **6-12m** | 1.7±83.6 | 791±2847 | 0.0822 |
| ***p* values** | <0.0001 | 0.0056 |  |

Quantification of the total expression of *L. donovani* mRNA in blood from P (ToD: n=12, EoT: n=16, 3m: n=8, 6-12m: n=7) and R VL/HIV patients (ToD: n=23, EoT: n=14, 3m: n=16, 6-12m: n=18)
Statistical differences between P and R VL/HIV patients at each time point were determined using a Mann-Whitney test; statistical differences between the 4 different time points for each cohort of patients were determined by Kruskal-Wallis test.

**Figure 3:**
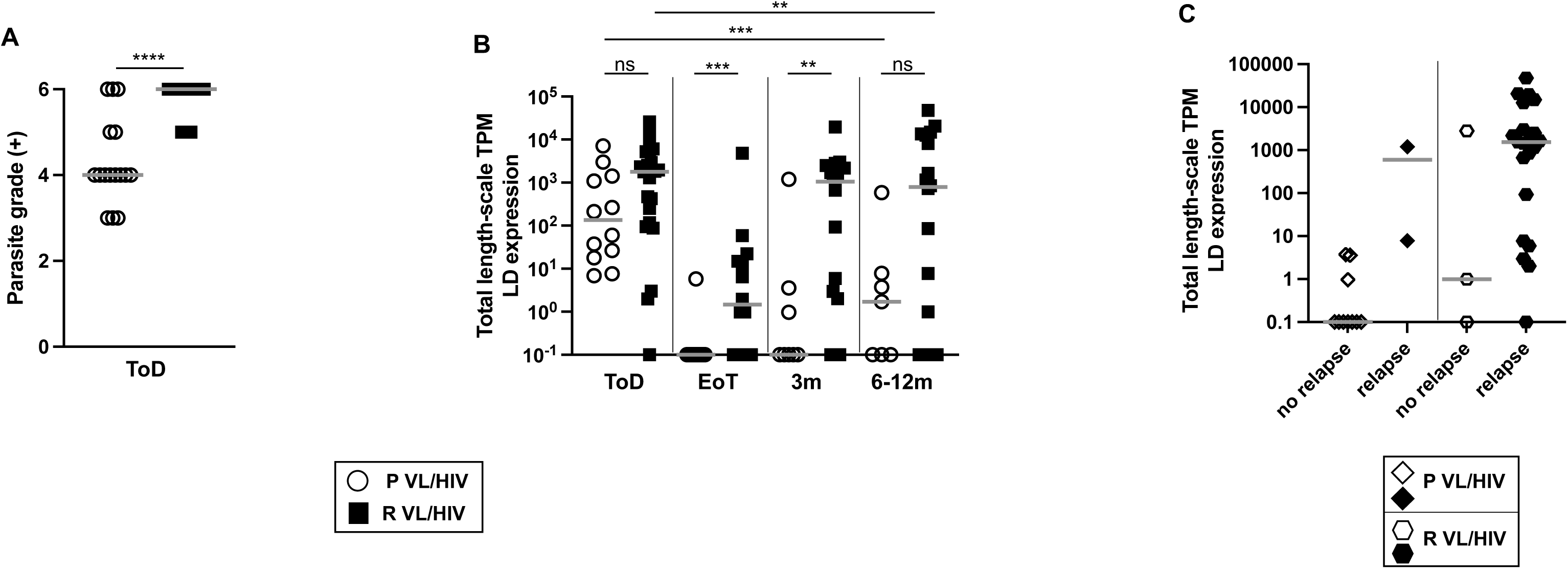
Parasite load: **A.** Quantification of *Leishmania* amastigotes in smears of splenic aspirates collected from P VL/HIV (n=16) and R VL/HIV (n=26) patients at ToD. **B.** Quantification of the total expression of *L. donovani* mRNA in blood from P (ToD: n=12, EoT: n=16, 3m: n=8, 6-12m: n=7) and R VL/HIVVL/HIV (ToD: n=23, EoT: n=14, 3m: n=16, 6-12m: n=18). **C.** Comparison of the total expression of *L. donovani* mRNA in blood from P VL/HIV who did not relapse (n=10) and those who did relapse (n=2) and R VL/HIV who did not relapse (n=21) and those who did relapse (n=3) after successful antileishmanial treatment during the 3- and 6-12-month follow-up period. If a patient did not relapse during the 2 time points of follow-up and if a patient relapsed at both 3, 6-12 months, this is represented as 2 measurements. Each symbol represents the value for one individual, the straight lines represent the median. Statistical differences between P and R VL/HIV patients (A) at each time point (B) were determined using a Mann-Whitney test; statistical differences between the 4 different time points (B) for each cohort of patients were determined by Kruskal-Wallis test. LD mRNA= *L. donovani* mRNA. ToD=Time of Diagnosis; EoT=End of Treatment; 3m=3 months post EoT; 6-12m=6-12 months post EoT.

Despite being on ART, the majority of patients with P VL/HIV and just under half of the R VL/HIV cohort had detectable plasma viral load (Table 5A). There were no significant differences in plasma viral load (Table 5B) between P and R VL/HIV patients at each time point and over time. No significant differences in viral load were observed between P and R VL/HIV who relapsed and those who didn’t relapse during follow-up (data not shown).

**Table 5A:** % of VL/HIV patients with detectable viral load.

|  | **P VL/HIV** | **R VL/HIV** |
| --- | --- | --- |
| **ToD** | 66.7% | 47.6% |
| **EoT** | 80.0% | 44.4% |
| **3m** | 27.3% | 50% |
| **6-12m** | 66.7% | 38.1% |

**Table 5B:** HIV-1 viral load in plasma (copies/ml of plasma)

|  | **P VL/HIV** | **R VL/HIV** | ***p* values** |
| --- | --- | --- | --- |
| **ToD** | 381±553679 | 200±581955 | 0.8452 |
| **EoT** | 150±50305 | 0.1±208331 | 0.4946 |
| **3m** | 542.5±160.9 | 2,873±99923 | 0.0799 |
| **6-12m** | 1,341±136928 | 629.5±199439 | 0.2758 |

**A.** % of P VL/HIV (ToD: n=18, EoT: n=15, 3m: n=11, 6-12m: n=6) and R VL/HIV (ToD: n=21, EoT: n=18, 3m: n=16, 6-12m: n=6) patients who had detectable viral loads at ToD, EoT, 3 and 6-12m. **B.** HIV-1 viral load in plasma from P VL/HIV and R VL/HIV patients. Statistical difference between P and R VL/HIV was determined by Mann-Whitney test. ToD=Time of Diagnosis; EoT=End of Treatment; 3m=3 months post EoT; 6-12m=6-12 months post EoT.

#### Clinical presentations

Next, we assessed whether P and R VL/HIV patients presented with different clinical manifestations. At ToD, fever was significantly higher in P VL/HIV patients as compared to R VL/HIV patients (Figure 4A) but normalised thereafter to levels similar to controls (controls: 36.0±0.1°C, *p*>0.05, data not shown). No significant differences in fever were observed between P and R VL/HIV patients who relapsed and those who didn’t relapse during follow-up (data not shown).

**Figure 4:**
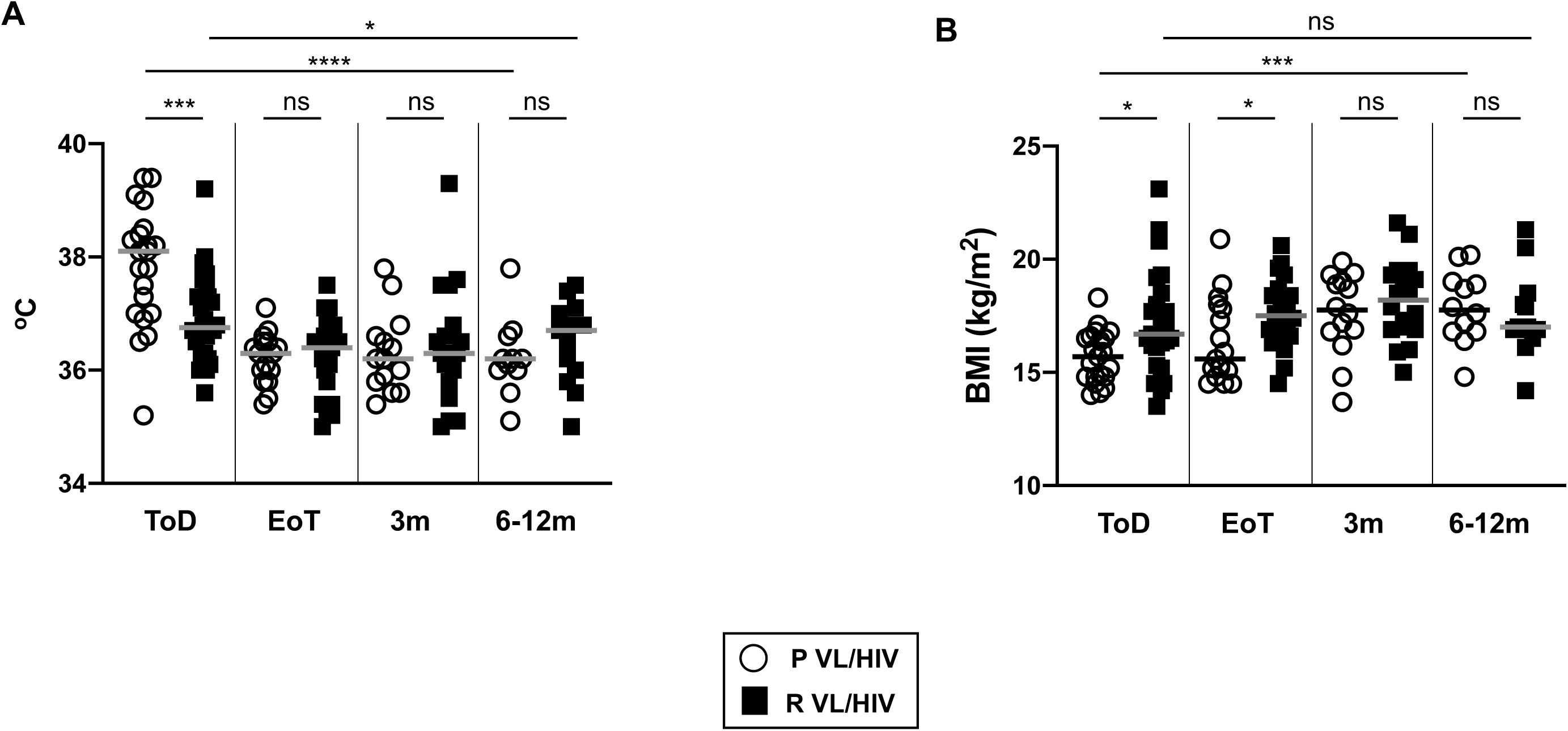
Clinical parameter: **A.** Body temperature was measured in P VL/HIV (ToD: n=21, EoT: n=16, 3m: n=14, 6-12m: n=11) and R VL/HIV (ToD: n=28, EoT: n=22, 3m: n=18, 6-12m: n=13) patients. **B.** BMI was calculated for P VL/HIV (ToD: n=21, EoT: n=17, 3m: n=14, 6-12m: n=12) and R VL/HIV (ToD: n=28, EoT: n=22, 3m: n=18, 6-12m: n=13) patients. Each symbol represents the value for one individual, the straight lines represent the median. Statistical differences between P VL/HIV and R VL/HIV patients at each time point were determined using a Mann-Whitney test; statistical differences between the 4 different time points for each cohort of patients were determined by Kruskal-Wallis test. ToD=Time of Diagnosis; EoT=End of Treatment; 3m=3 months post EoT; 6-12m=6-12 months post EoT. ns=not significant.

The median BMI of each group of VL/HIV patients was below the normal value of 18.5 (34) throughout follow-up and was significantly higher in R than P VL/HIV at ToD and EoT (*p*=0.0173 and *p*=0.0192 respectively, Figure 4B). The BMI of P VL/HIV, but not R VL/HIV patients, increased significantly over time (*p*=0.0010). No significant differences in BMI were observed between P and R VL/HIV patients who relapsed and those who didn’t relapse during follow-up (data not shown).

Since hepatosplenomegaly is a typical sign of VL/HIV patients, spleen and liver sizes as measured below the costal margin are systematically recorded when patients present to the clinic. As shown in Figure 5A, spleen sizes were similar in both groups at ToD, EoT and 3m and decreased throughout follow-up, but were significantly higher at 6-12m in R VL/HIV (*p*=0.0061). To assess whether this increase in spleen size was due to VL relapse, we compared the spleen size of patients who didn’t relapse with those who relapsed after initial cure in P and R VL/HIV groups. As shown in Figure 5B, there was no significant difference in spleen sizes between the patients who did not relapse during follow-up in both cohorts (P and R VL/HIV patients). As expected, the spleen sizes were significantly higher in P and R VL/HIV patients who relapsed during follow-up in both groups (Figure 5B).

**Figure 5:**
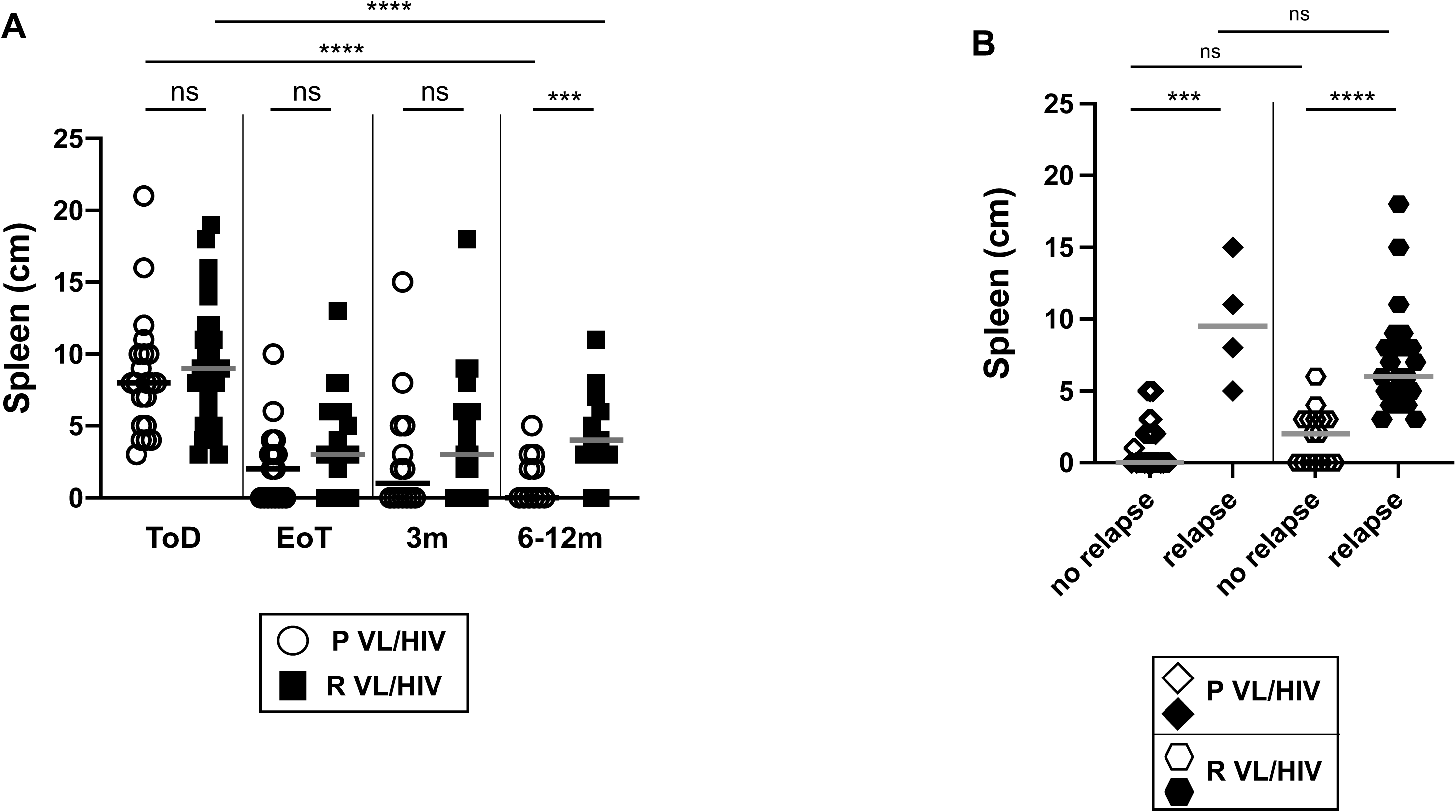
**A.** Spleen size as measured in cm below the costal margin on VL (ToD: n=21, EoT: n=17, 3m: n=14, 6-12m: n=11), VL/HIV (ToD: n=28, EoT: n=22, 3m: n=18, 6-12m: n=13) patients. **B.** Comparison of the spleen of P VL/HIV who did not relapse (n=25) and those who did relapse (n=4) and R VL/HIV who did not relapse (n=16) and those who did relapse (n=23) after successful antileishmanial treatment during the 3- and 6-12-month follow-up period. If a patient did not relapse during the 2 time points of follow-up and if a patient relapsed at both 3, 6-12 months, this is represented as 2 measurements. Each symbol represents the value for one individual, the straight lines represent the median. Statistical differences between 2 groups were determined using a Mann-Whitney test; statistical differences between the 4 different time points for each cohort of patients were determined by Kruskal-Wallis test. ToD=Time of Diagnosis; EoT=End of Treatment; 3m=3 months post EoT; 6-12m=6-12 months post EoT. ns=not significant.

The liver was also clearly measurable below the costal margin at ToD in both groups of patients and decreased throughout follow-up (Table 6). No significant differences in liver sizes were observed between P and R VL/HIV who relapsed and those who didn’t relapse during follow-up (data not shown).

**Table 6:** Liver size.

|  | **P VL/HIV** | **R VL/HIV** | ***p* values** |
| --- | --- | --- | --- |
| **ToD** | 4.0±0.6 | 2.0±0.7 | 0.2191 |
| **EoT** | 0.0±0.2 | 0.0±0.4 | 0.7136 |
| **3m** | 0.0±0.3 | 0.0±0.4 | 0.9263 |
| **6-12m** | 0.0±0.3 | 0.0±0.3 | 0.6133 |
| ***p* values** | <0.0001 | 0.009 |  |

Liver size was measured in cm below the costal margin on P VL/HIV (ToD: n=21, EoT: n=17, 3m: n=14, 6-12m: n=18) and R VL/HIV (ToD: n=28, EoT: n=22, 3m: n=18, 6-12m: n=22) patients.
Statistical differences between P VL/HIV and R VL/HIV patients at each time point were determined using a Mann-Whitney test and statistical differences between the 4 different time points for each cohort of patients were determined by Kruskal-Wallis test.
ToD=Time of Diagnosis; EoT=End of Treatment; 3m=3 months post EoT; 6-12m=6-12 months post EoT. ns=not significant.

#### Haematological profiles

White blood cell counts (WBCs) were similar in both groups of VL/HIV patients at ToD and increased at EoT, however, at 6-12m R VL/HIV patients had significantly lower WBCs than P VL/HIV patients (Table 7A). RBCs increased significantly in P VL/HIV patients until the 3m time point and plateaued at 6-12m; however, no significant improvement in RBCs was observed in the blood of R VL/HIV patients throughout the follow-up (Table 7B). Similarly, platelet (PLT) counts increased at EoT in P VL/HIV patients, plateaued thereafter and were higher than in R VL/HIV patients at 3 and 6-12 months (Table 7C); PLT counts of R VL/HIV patients did not change significantly throughout the follow up (Table 7C). Despite an increase in WBC, RBC and PLT counts in the blood of P VL/HIV, these values still remained significantly lower than those of healthy controls (p<0.0001, data not shown).

**Table 7A:** White blood cell counts (WBC, cells/μl of blood)

| WBCs | P VL/HIV | R VL/HIV | *p* values |
| --- | --- | --- | --- |
| ToD | 1,500±165 | 1,900±374 | 0.0659 |
| EoT | 4,100±325 | 3,100±440 | 0.8399 |
| 3m | 4,350±514 | 3,485±307 | 0.0606 |
| 6-12m | 3,800±613 | 2,450±338 | 0.0388 |
| *p* values | <0.0001 | 0.0162 |  |

**Table 7B:**
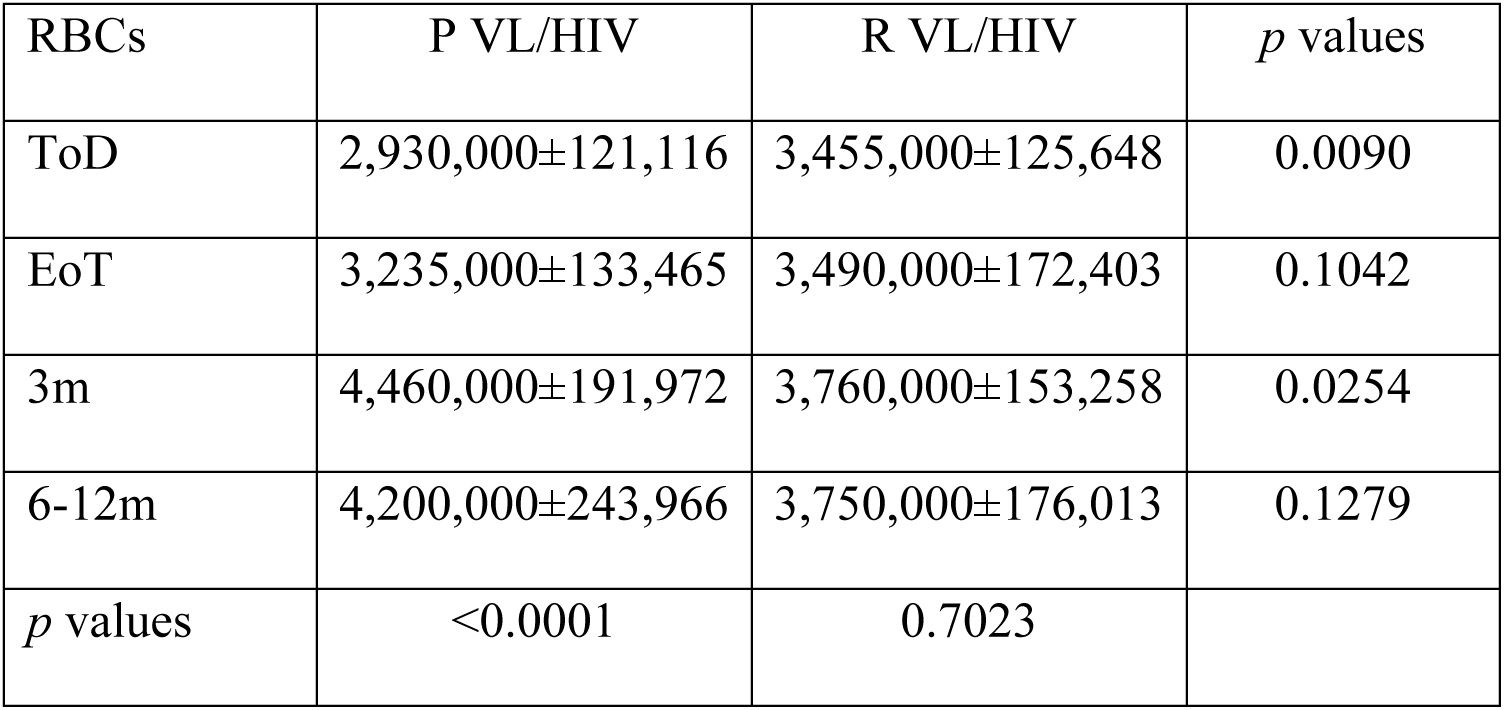
Red blood cell counts (RBC, cells/μl of blood)

| RBCs | P VL/HIV | R VL/HIV | *p* values |
| --- | --- | --- | --- |
| ToD | 2,930,000±121,116 | 3,455,000±125,648 | 0.0090 |
| EoT | 3,235,000±133,465 | 3,490,000±172,403 | 0.1042 |
| 3m | 4,460,000±191,972 | 3,760,000±153,258 | 0.0254 |
| 6-12m | 4,200,000±243,966 | 3,750,000±176,013 | 0.1279 |
| *p* values | <0.0001 | 0.7023 |  |

**Table 7C:**
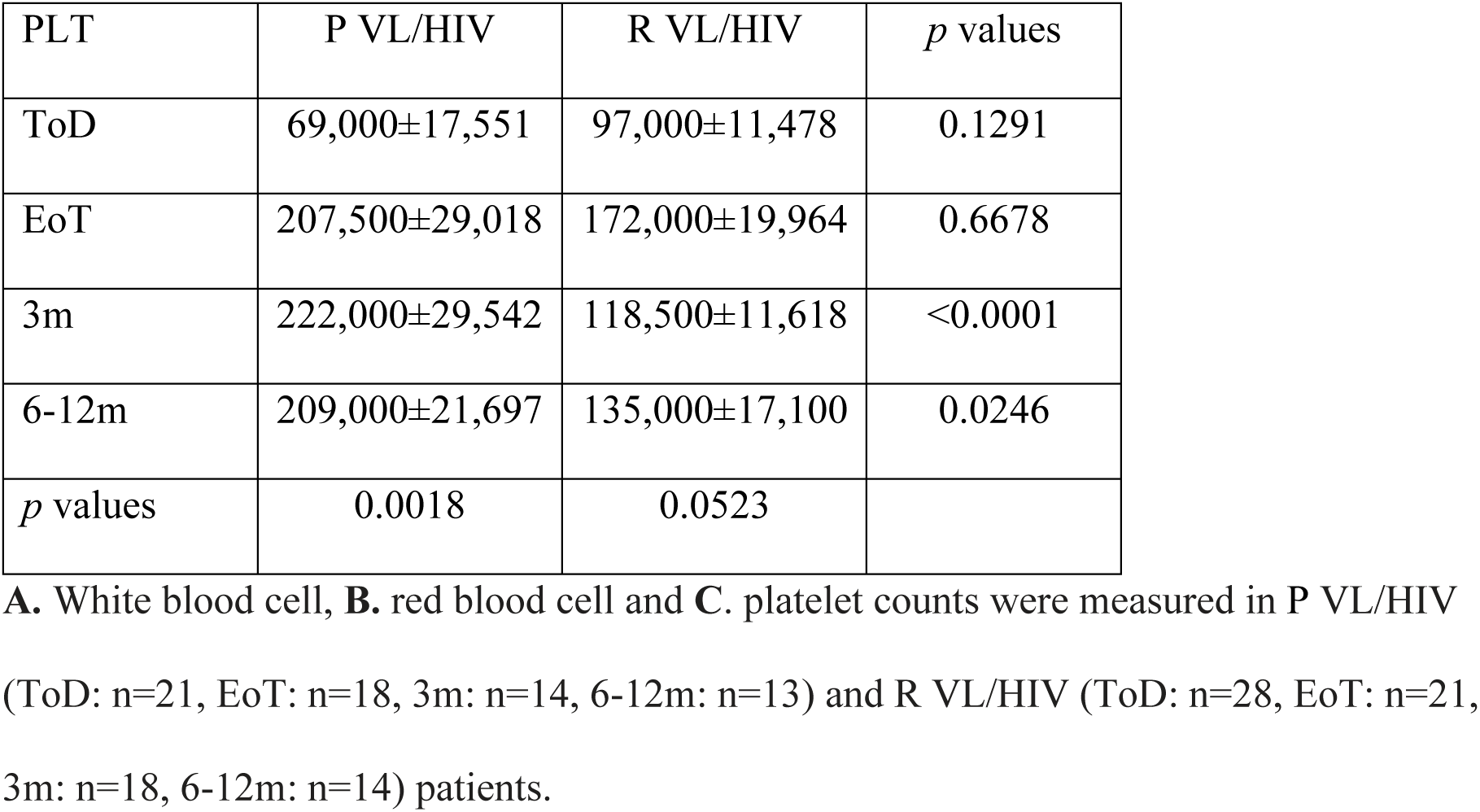
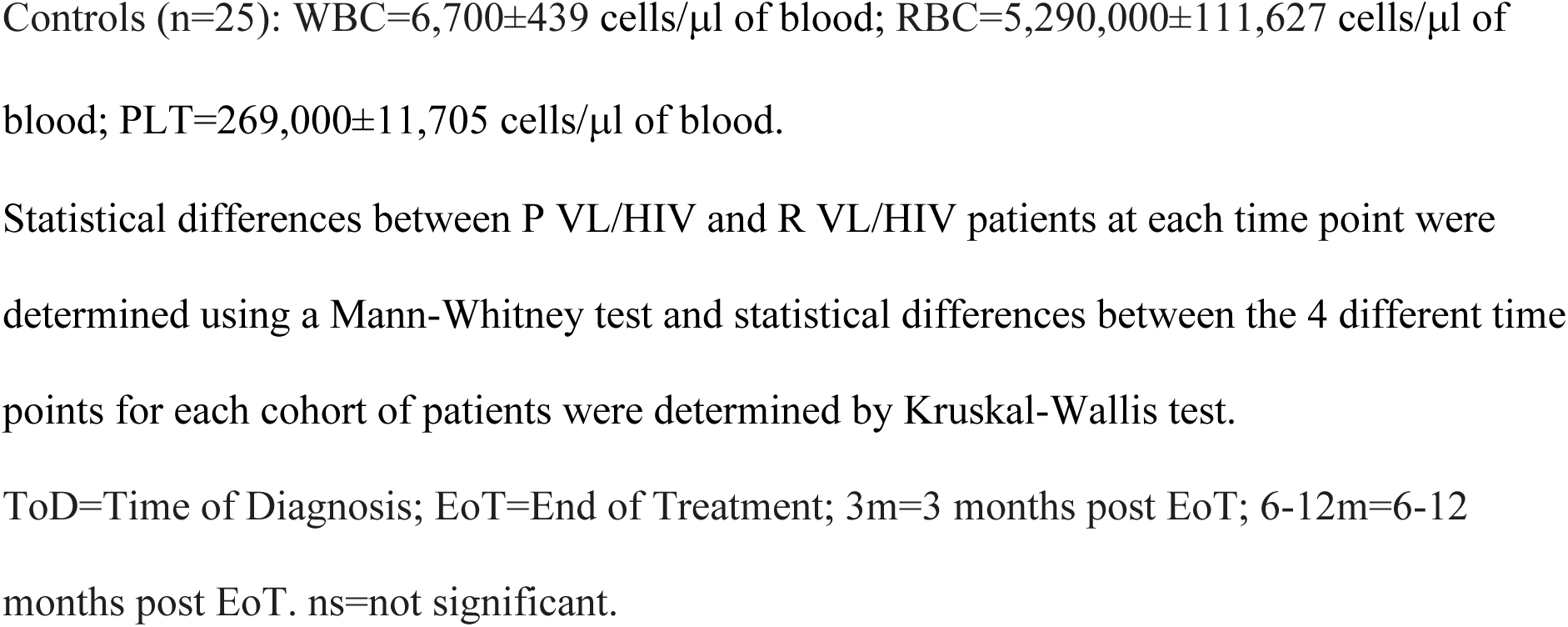
Platelet counts (PLT, cells/μl of blood)

#### Antigen-specific production of IFNγ and IL-10 by whole blood cells from P and R VL/HIV patients

One of the hallmarks of VL/HIV patients is the inefficiency of whole blood cells to produce IFNγ in response to *Leishmania*-specific stimulation at time of diagnosis and throughout follow-up (12). Here we compared *Leishmania*-specific IFNγ production by whole blood cells from P and R VL/HIV patients. Results presented in Figure 6A show that whereas the levels of IFNγ did not significantly change over time in both groups, whole blood cells from R VL/HIV patients produced significantly less IFNγ than whole blood cells from P VL/HIV patients throughout follow-up. Next, we assessed whether whole blood cells from P and R VL/HIV patients who didn’t relapse after initial cure produced more IFNγ. As shown in Figure 6B, there was more *Leishmania*-specific IFNγ produced by whole blood cells from P VL/HIV who did not relapse as compared to R VL/HIV patients who did not relapse; however, blood was collected from 2 out of the 4 P VL/HIV patients who relapsed during follow-up, it was therefore not possible to draw meaningful conclusions from these data. In the R VL/HIV group, the levels of IFNγ were significantly higher in patients who had not relapsed during follow-up (Figure 6B). The production of IFNγ in response to PHA remained similar in both groups throughout follow-up (Table 8A).

**Figure 6:**
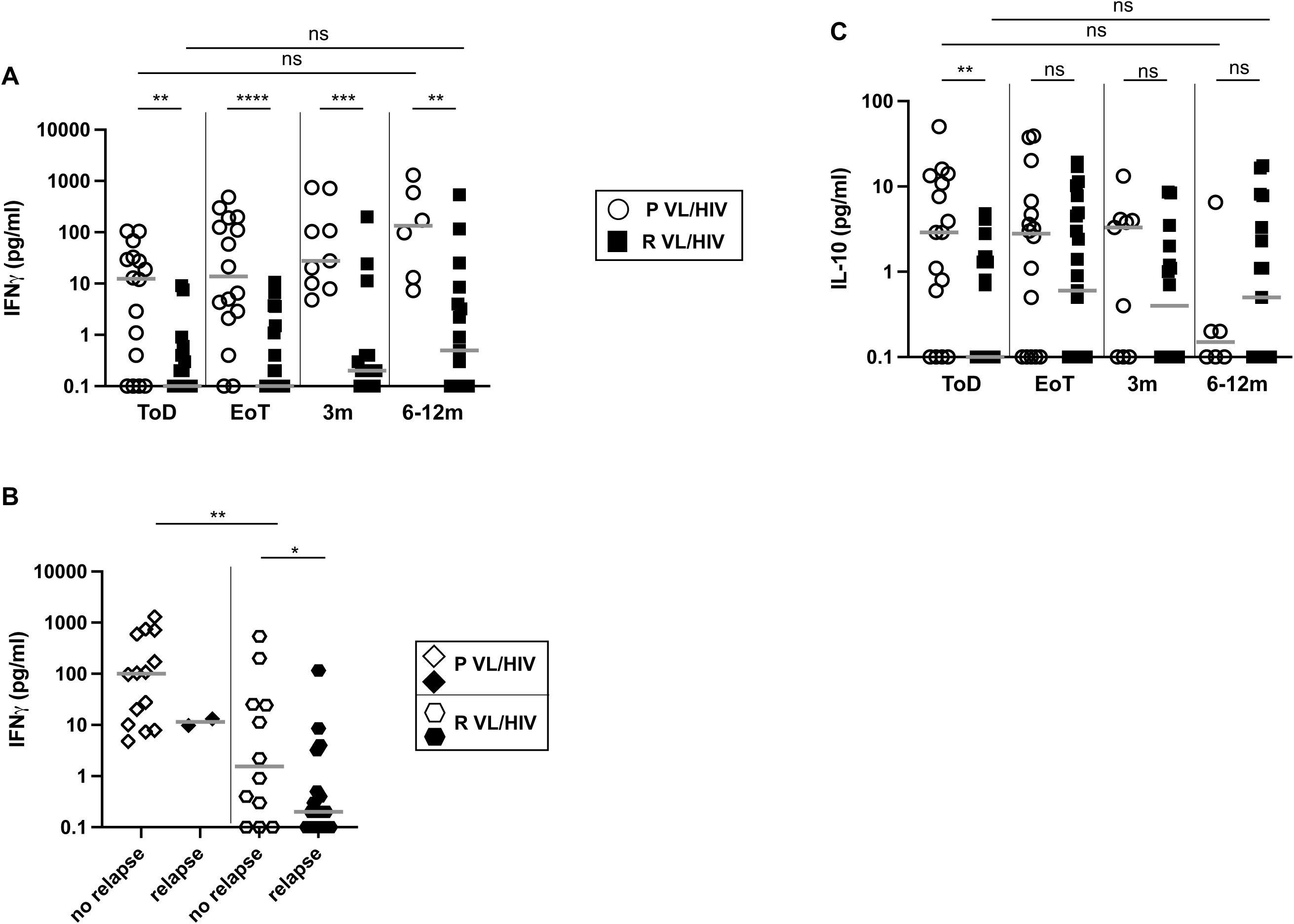
Whole blood assay: antigen-specific production of IFNγ and IL-10. Whole blood cells from P VL/HIV (ToD: n=16, EoT: n=16, 3m: n=9, 6-12m: n=6) and R VL/HIV (ToD: n=22, EoT: n=23, 3m: n=16, 6-12m: n=17) patients were cultured in the presence of SLA and PBS. **A**. IFNγ and **C**. IL-10 levels in the supernatant were measured by ELISA after 24hrs and the values obtained with PBS alone was deducted from the value obtained with SLA. **B.** Comparison of IFNγ levels in the supernatant of whole blood cells from P VL/HIV who did not relapse (n=14) and those who did relapse (n=2) and R VL/HIV who did not relapse (n=12) and those who did relapse (n=21) after successful antileishmanial treatment during the 3- and 6-12-month follow-up period. If a patient did not relapse during the 2 time points of follow-up and if a patient relapsed at both 3, 6-12 months, this is represented as 2 measurements. Statistical differences between two groups were determined using a Mann-Whitney test; statistical differences between the 4 different time points for each cohort of patients were determined by Kruskal-Wallis test. ToD=Time of Diagnosis; EoT=End of Treatment; 3m=3 months post EoT; 6-12m=6-12 months post EoT.

**Table 8A:** Production of IFNγ in response to PHA.

| IFNγ (pg/ml) | P VL/HIV | R VL/HIV | *p* values |
| --- | --- | --- | --- |
| **ToD** | 10.5±12.0 | 5.7±114.4 | 0.7534 |
| **EoT** | 24.6±143.7 | 39.1±197.2 | 0.4746 |
| **3m** | 55.5±170.9 | 5.3±126.1 | 0.1476 |
| **6-12m** | 30.4±36.2 | 15.8±209.7 | 0.8002 |
| ***p* values** | 0.3327 | 0.5585 |  |

We have previously shown that *Leishmania*-specific production of IL-10 is impaired in the WBA in VL/HIV patients (12) and that it was not associated with disease severity. Our results presented in Figure 6C show that as compared to P VL/HIV patients, antigen-specific production of IL-10 by R VL/HIV whole blood cells was significantly lower at ToD but similar at EoT, 3 and 6-12m. No significant differences in antigen-specific IL-10 were observed between P and R VL/HIV who relapsed and those who didn’t relapse during follow-up (data not shown).

The production of IL-10 in response to PHA increased significantly over time in both groups but was lower at 6-12m in R VL/HIV patients (Table 8B).

**Table 8B:** Production of IL-10 in response to PHA.

| IL-10 (pg/ml) | P VL/HIV | R VL/HIV | *p* values |
| --- | --- | --- | --- |
| **ToD** | 17.1±13.1 | 30.0±28.5 | 0.6089 |
| **EoT** | 145.8 ±49.3 | 224.9±54.8 | 0.5298 |
| **3m** | 360.0±62.7 | 306.9±56.1 | >0.9999 |
| **6-12m** | 486.7±147.0 | 131.8±43.2 | 0.0126 |
| ***p* values** | <0.0001 | 0.008 |  |

Whole blood cells from P VL/HIV (ToD: n=16, EoT: n=16, 3m: n=9, 6-12m: n=6) and R VL/HIV (ToD: n=22, EoT: n=23, 3m: n=16, 6-12m: n=17) patients were cultured in the presence of d PHA. **A.** IFNγ and **B.** IL-10 levels in the supernatant were measured by ELISA after 24hrs.
Statistical differences between P and R VL/HIV patients at each time point were determined using a Mann-Whitney test; statistical differences between the 4 different time points for each cohort of patients were determined by Kruskal-Wallis test.
ToD=Time of Diagnosis; EoT=End of Treatment; 3m=3 months post EoT; 6-12m=6-12 months post EoT.

#### CD4^+^ and CD8^+^ T cell counts and PD1 expression

Our previous results showed that in VL/HIV patients, the failure to restore antigen-specific production of IFNγ correlated with persistently low CD4^+^ T cell counts and high expression of PD1 on CD4^+^ T cells (12). Results presented in Figure 7A show that despite low CD4^+^ T cell counts in both P and R VL/HIV patients throughout follow-up as compared to controls (*p*<0.0001), CD4^+^ T cell counts increased significantly in the P VL/HIV group, but not in the R VL/HIV group (Figure 7A). To assess whether P and R VL/HIV patients who didn’t relapse after clinical cure had higher CD4^+^ T cell counts, the 2 cohorts were subdivided into patients who relapsed and those who didn’t. Results presented in Figure 7B show that P VL/HIV patients who didn’t relapse had significantly higher CD4^+^ T cell counts as compared to R VL/HIV. In the P VL/HIV group who relapsed, data were collected for three patients, it was therefore not possible to draw meaningful conclusions from these results. In contrast, CD8^+^ T cell counts were similar between both groups of VL/HIV patients at all time points and were restored to levels similar to those of controls at EoT (*p*>0.05). No significant differences in CD8^+^ T cell counts were observed between P and R VL/HIV who relapsed and those who didn’t relapse during follow-up (data not shown).

**Figure 7:**
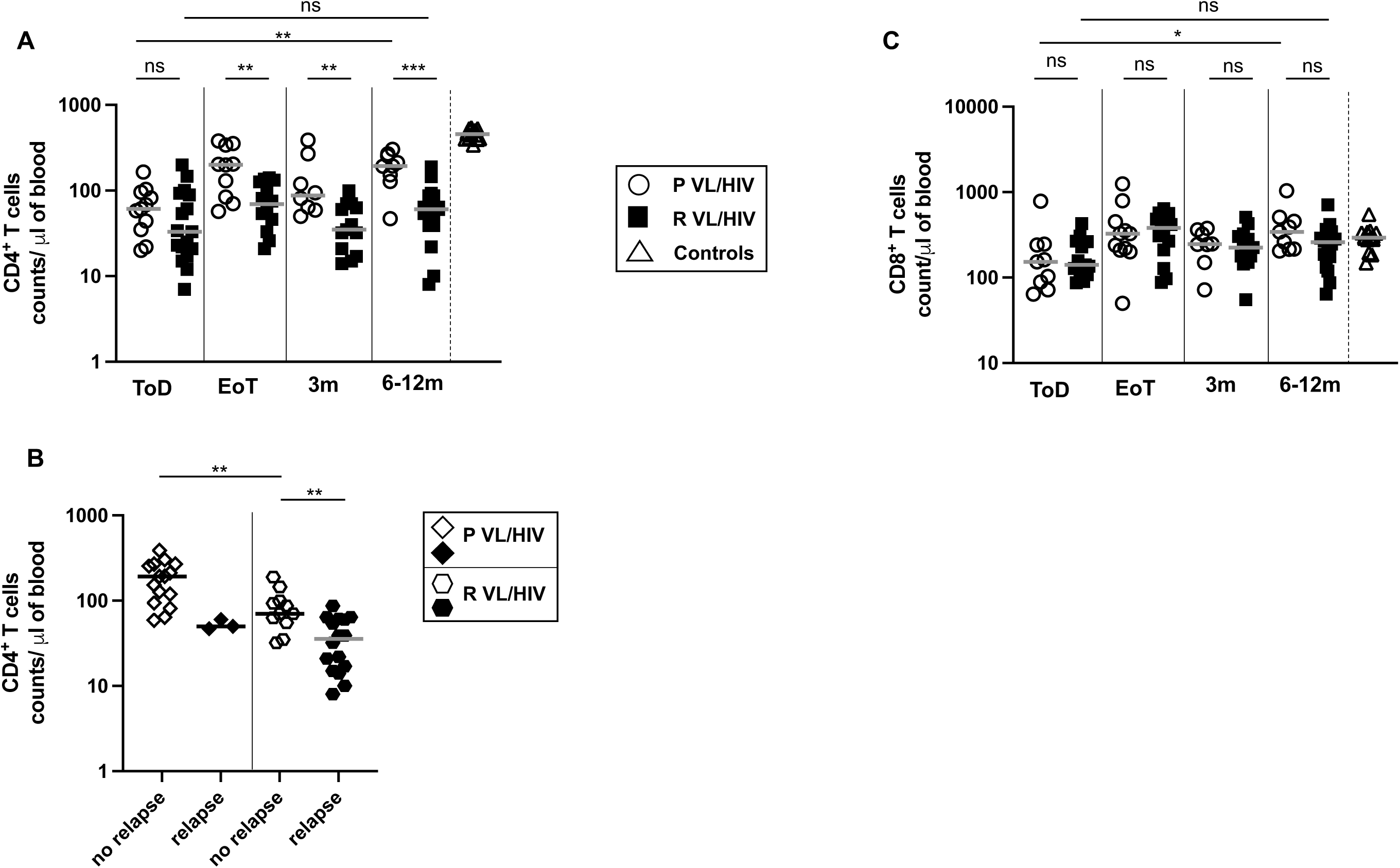
CD4^+^ and CD8^+^ T cell counts: **A.** CD4^+^ T cell counts in the blood of P VL/HIV (ToD: n=11, EoT: n=10; 3m: n=8, 6-12m: n=9) and R VL/HIV (ToD: n=17, EoT: n=14, 3m: n=13, 6-12m: n=16) patients; and **C.** CD8^+^ T cell counts in the blood of P VL/HIV (ToD: n=13, EoT: n=12, 3m: n=8, 6-12m: n=6) and R VL/HIV (ToD: n=11, EoT: n=12, 3m: n=13, 6-12m: n=10) patients were measured by flow cytometry. **B.** Comparison of CD4^+^ T cell counts in the blood of P VL/HIV who did not relapse (n=15) and those who did relapse (n=3) and R VL/HIV who did not relapse (n=11) and those who did relapse (n=16) after successful antileishmanial treatment during the 3- and 6-12-month follow-up period. If a patient did not relapse during the 2 time points of follow-up and if a patient relapsed at both 3, 6-12 months, this is represented as 2 measurements. Statistical differences between two groups were determined using a Mann-Whitney test; statistical differences between the 4 different time points for each cohort of patients were determined by Kruskal-Wallis test. ToD=Time of Diagnosis; EoT=End of Treatment; 3m=3 months post EoT; 6-12m=6-12 months post EoT.

We have previously shown that the expression level of PD1, an inhibitory receptor that can be associated with impaired effector functions, remained high on CD4^+^ T cells in VL/HIV patients (12). Results presented in Figure 8A show that CD4 PD1 iMFI were higher throughout follow-up in both P and R VL/HIV patients as compared to controls (*p*<0.0001), and that CD4 PD1 iMFI levels decreased significantly in the P VL/HIV group, but not in the R VL/HIV group. There was no significant difference between the medians of CD4 PD1 iMFI in P and R VL/HIV patients who didn’t relpase over time (Figure 8B). CD4 PD1 iMFI was significantly higher in R VL/HIV who relapsed as compare to those who didn’t relapse (Figure 8B). In the P VL/HIV group who relapsed, data were collected for three patients, it was therefore not possible to draw meaningful conclusions from these results.

**Figure 8:**
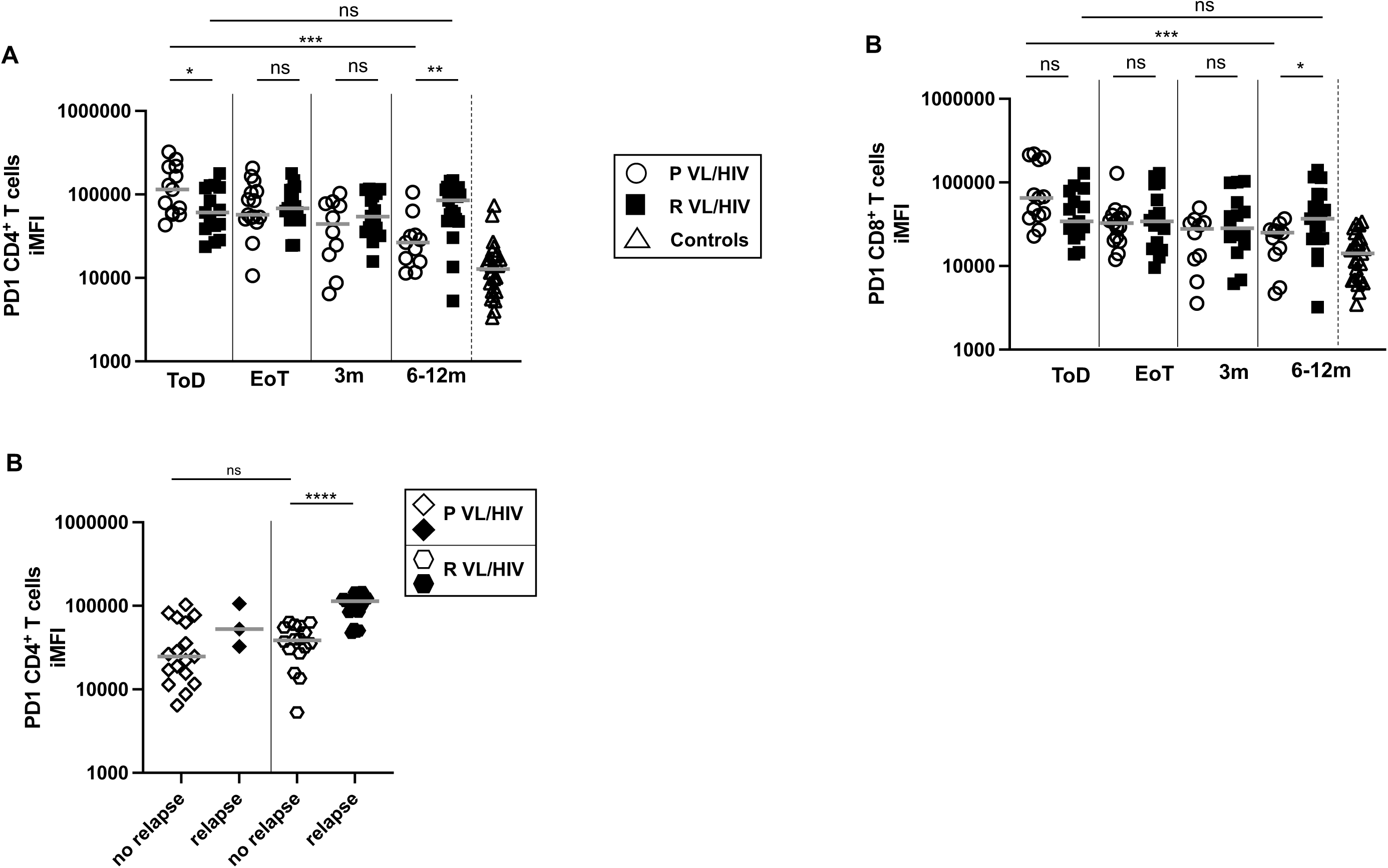
PD1 expression on CD4^+^ and CD8^+^ T cells: **A.** CD4 PD1 iMFI and **C.** CD8 PD1 iMFI was measured by multiplying the % of T cells and the median fluorescence intensity of PD1 as measured by flow cytometry in the PBMCs of P VL/HIV (ToD: n=13, EoT: n=15, 3m: n=10, 6-12m: n=11), R VL/HIV patients (ToD: n=15, EoT: n=17, 3m: n=16, 6-12m: n=21) and healthy controls (n=10). **B.** Comparison of CD4 PD1 iMFI in P VL/HIV who did not relapse (n=17) and those who did relapse (n=3) and R VL/HIV who did not relapse (n=16) and those who did relapse (n=21) after successful antileishmanial treatment during the 3- and 6-12-month follow-up period. If a patient did not relapse during the 2 time points of follow-up and if a patient relapsed at both 3, 6-12 months, this is represented as 2 measurements. Statistical differences between two groups were determined using a Mann-Whitney test; statistical differences between the 4 different time points for each cohort of patients were determined by Kruskal-Wallis test. ToD=Time of Diagnosis; EoT=End of Treatment; 3m=3 months post EoT; 6-12m=6-12 months post EoT.

Although CD8 PD1 MFI remained significantly higher in both VL/HIV groups as compared to controls throughout follow-up (Figure 8C, *p*<0.0001), CD8 PD1 iMFI decreased significantly over time in P VL/HIV, but not R VL/HIV. No significant differences in CD8 PD1 iMFI were observed between P and R VL/HIV who relapsed and those who didn’t relapse during follow-up (data not shown).

#### Survival analysis

Next, we tested the association between clinical and immunological factors that could be measured at time of diagnosis or at the end of treatment and the rate of VL relapse; reasoning that identifying associations at these early timepoints could help identify patients who could most benefit from additional intervention to prevent relapse. As shown in Table 9, prior history of VL relapse (recurrent VL) is an important risk factor for future relapse, with an estimated 1-year relapse rate of almost 79% (CI 49.7-91.9) in recurrent VL patients compared to only 22.5% in patients experiencing their primary episode of VL (CI 0.3%-39.7%). In an effort to understand the causes of this further relapse, we tested the association of a wide range of clinical and immunological factors measured at both time of diagnosis and end of treatment for association with relapse rates, and found only two other factors showing significant associations – CD4^+^ T cell count at end of treatment and parasite load measured by splenic aspirate at time of diagnosis. Neither of these factors remained significant in two-variable models that also included prior VL history, while VL history remained independently associated with relapse rate in these models.

**Table 9.**
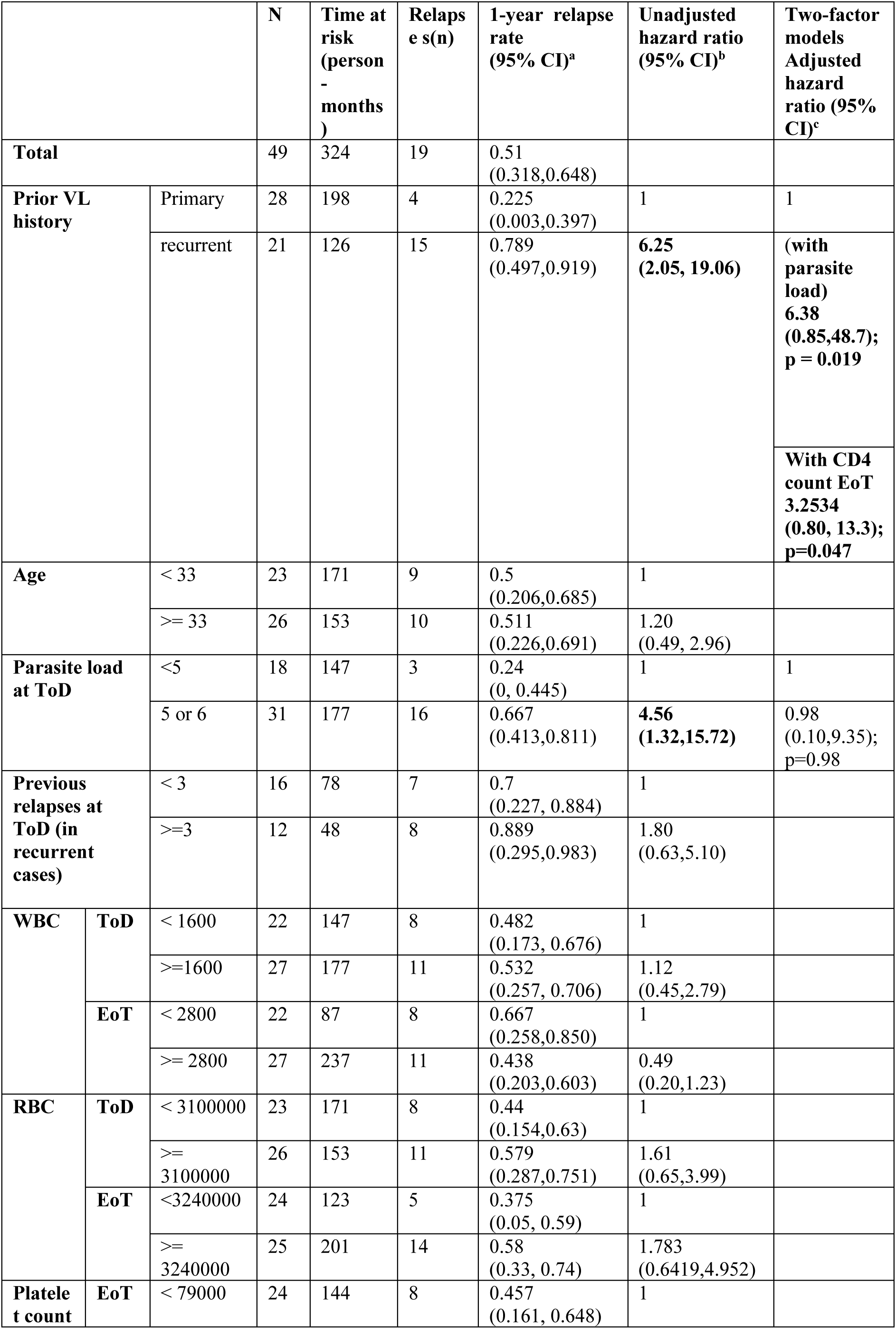
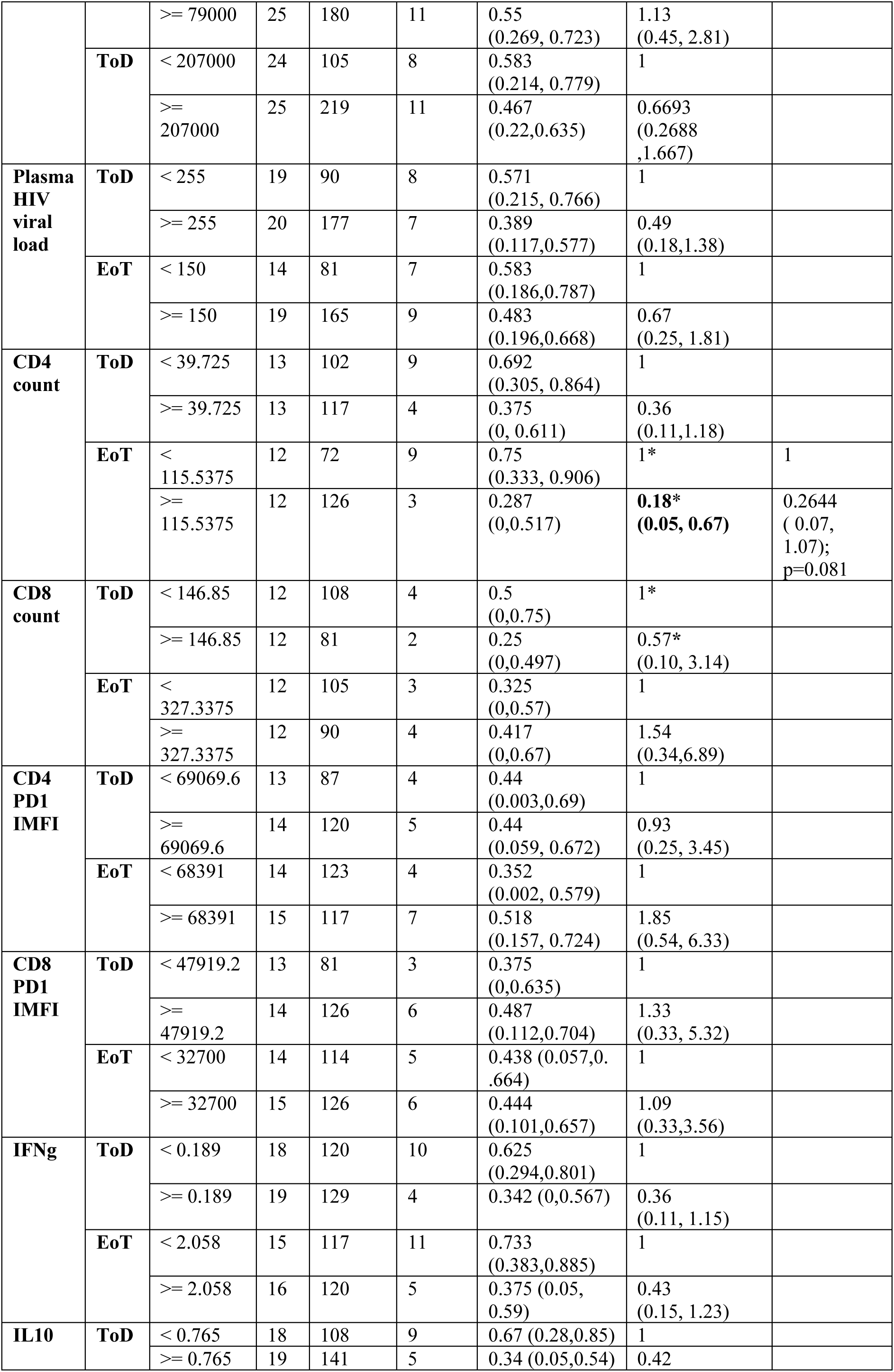
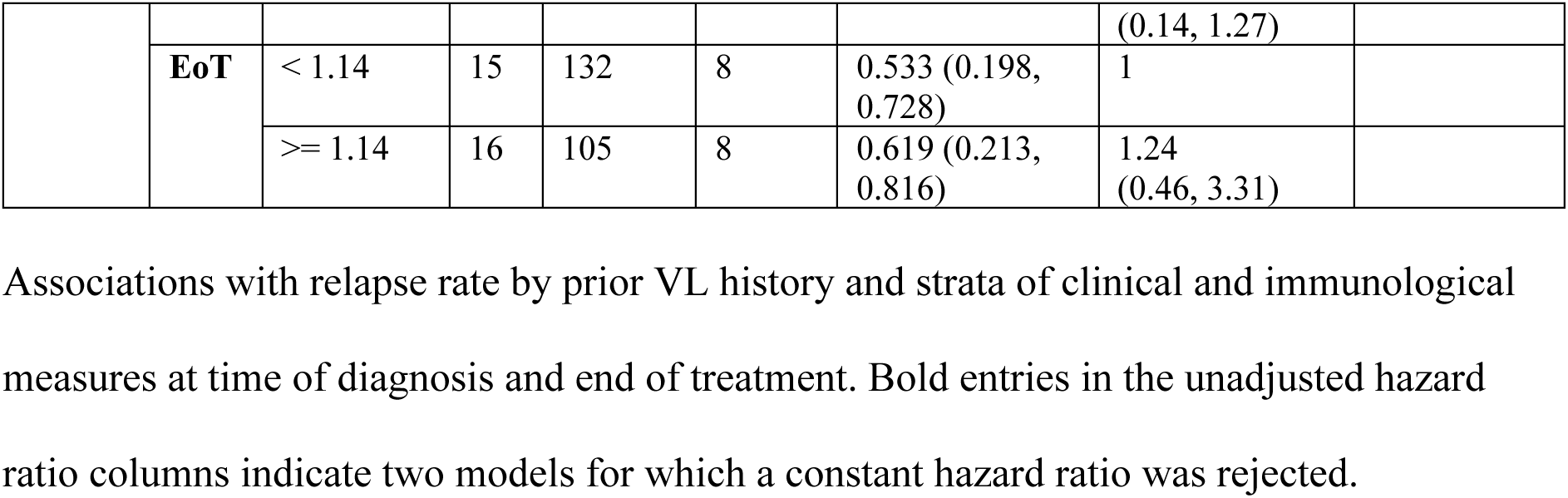
Survival analysis.

## DISCUSSION

We have previously shown that over a period of 3 years, VL/HIV patients experience a high rate of relapse (12). Here we analysed this cohort of patients further by comparing clinical and immunological parameters in VL/HIV patients who presented with their first episode of VL and those with a previous history of VL relapse.

Our results show that there was a poorer recovery in weight gain and blood cell counts, higher spleen size and parasite load in R VL/HIV patients, as compared to P VL/HIV patients:

- The median BMI of both groups of patients remained below 18.5 throughout the follow-up, however the BMI increased significantly in the P, but not the R VL/HIV cohort. Malnutrition plays a crucial role in increased susceptibility to infection and/or disease severity by weakening both innate and acquired immunity (35, 36). It is therefore possible that the lower BMI observed in R VL/HIV patients at 6-12 months contributes to their poorer prognosis. Better management of malnutrition in both groups of VL/HIV patients could improve their ability to mount an effective immune response.
- Splenomegaly increased in R VL/HIV patients during the follow-up period, consistent with the observed increase in the parasite load over time in this group of patients.
- Although both groups of patients remained pancytopenic, the increase in WBC, RBC and PLT counts following treatment was less efficient in R VL/HIV patients. Bone marrow suppression can contribute to pancytopenia, and both VL and HIV infection have been associated with bone marrow failure (37–39). Both pathogens can infect hematopoietic stem/progenitor cells (HSPCs), and this can impair haematopoiesis. The higher parasite load observed in R VL/HIV patients might contribute to the poorer recovery of all blood cell lineages.
- Although the parasite load decreased at EoT, the load as measured in blood remained higher from EoT onwards in R VL/HIV patients; at 3 and 6-12m it was similar to the load at ToD. These results are consistent with the higher relapse rate in these patients, 85% in the R VL/HIV versus 65% in the P VL/HIV cohort, indicating that R VL/HIV patients have a poorer ability to control parasite replication.

To identify possible mechanisms responsible for this inability to efficiently control parasite replication, we analysed immunological parameters. It has been shown that in splenic aspirates, IFNγ produced by CD4^+^ T cells contribute to parasite killing (40). We and others have speculated that in VL/HIV patients, the impairment of antigen-specific IFNγ production by CD4^+^ T cells plays a key role in the inefficient control of parasite replication (12, 40).

Here we show that the levels of antigen-specific IFNγ produced by whole blood cells were even further reduced in R VL/HIV as compared to P VL/HIV patients. This reduction in IFNγ might explain the higher parasite load detected in splenic aspirates and blood of R VL/HIV patients. Antigen-specific IFNγ is mainly produced by CD4^+^ T cells in the whole blood assay (40). It is therefore plausible that low IFNγ levels are a consequence of low CD4^+^ T cell counts. Here we show that both factors are likely to be involved: the CD4^+^ T cell counts are even lower in R VL/HIV and are accompanied with significantly lower production of antigen-specific IFNγ in the WBA, as compared to P VL/HIV.

Low CD4^+^ T cell counts could be due to poor HIV control: in our study, many patients still had detectable viral loads throughout the follow-up despite being on ART. The recovery of CD4^+^ cells is often stunted in individuals who started with low CD4^+^ T cell counts (41–43); but there is no ART regimen that has been shown to boost CD4^+^ T cell recovery. Inclusion of dolutegravir in first line treatment could help to improve CD4^+^ T cell recovery through more efficient suppression of viral replication (44). It is also possible that there is some resistance to HIV drugs in this population: this has been reported to both first and second-line treatments (45). Poor HIV control may also be due to difficulties accessing ART for the population of migrant workers during the agricultural season (12); indeed, this population is highly mobile and frequently lack access to health facilities where they can get ART. Poor adherence to ART is also likely to play a role: whereas adherence counselling is available to HIV patients, a recent study about ART adherence in the hospital in Gondar showed that adherence to ART was negatively associated with rural residence, lack of knowledge about HIV and ART, undisclosed HIV status to partners, and low CD4 count (46).

T cell exhaustion is another factor that might account for the low antigen-specific IFNγ level observed in VL/HIV patients. T cell exhaustion is characterised by a gradual loss of effector functions and co-expression of inhibitory receptors (47–49). PD1 is upregulated on T cells by signals such as IL-2, IL-7, type I IFNs and signalling via the TCR: it is therefore a marker of T cell activation (50). However, during chronic infection, the levels of PD1 remain high and are associated with T cell dysfunction (49). It is therefore plausible that in VL/HIV patients, persistent antigenic stimulation due to *Leishmania* and HIV contributes to T cell exhaustion. T cells responding to chronic infection undergo progressive loss of functions; since R VL/HIV patients have had a more intense and longer exposure to both pathogens, this could have resulted in higher PD1 CD4 iMFI and lower antigen-specific IFNγ production as compared to P VL/HIV. Our results also show that whereas CD8^+^ T cell counts are restored at EoT, CD8 PD1 iMFI decreased significantly in P VL/HIV but not in R VL/HIV patients, suggesting that CD8^+^ T cells may also have an exhausted phenotype. It has been previously shown that CD8^+^ T cells from the blood of VL patients have an anergic/exhausted phenotype, as shown by high levels of CTLA4 and PD1 (51). However, we did not detect CTLA4 on T cells in VL/HIV patients (12). In the study by Gautam *et al.*, the authors show that CD8^+^ T cells contribute to the basal levels of IFNγ in whole blood, but not to the antigen-specific IFNγ production (51); similarly the study by Kumar *et al.* showed that CD4^+^ T cells but not CD8^+^ T cells produce IFNγ in the WBA (40). We have previously discussed fundamental differences between VL patients in India and in Ethiopia: whereas whole blood cells from VL patients from Northwest Ethiopia have an impaired ability to produce antigen-specific IFNγ at ToD, this is not altered in VL patients from India (12, 31, 52). We hypothesised that these differences were associated with the less severe VL symptoms in Indian patients as compared to patients from Northwest Ethiopia (31). In view of these differences, since we did not determine the phenotype of IFNγ producing cells in our WBA, we cannot exclude the possibility that CD8^+^ T cells produce antigen-specific IFNγ. It is also possible that CD8^+^ T cells contribute to the elevated levels of IFNγ detected in the plasma of VL/HIV patients (12). VL/HIV patients are likely to play a major role in the transmission of VL. A recent study by Singh et al. showed that patients with active VL, but not asymptomatic or successfully treated VL patients, can transmit the parasites to sand flies (53). Another study showed that VL/HIV co-infected individuals transmitted the parasites most efficiently to the sand fly vectors (54). Since VL/HIV patients harbour higher parasite loads than VL patients, these co-infected individuals are likely to be a significant reservoir for *L. donovani* and have a high potential for parasite transmission; thereby preventing the elimination of visceral leishmaniasis. From a public health perspective, it is important to note not only the high parasite burden in these patients but also the potential for drug resistance to emerge. Given the importance of parasite load to transmission, the contribution of treatment failure in VL/HIV patients to the reservoir in the community needs to be determined and the cost to the health service as well as the health implications to the individual considered when determining management.

In summary our results show that VL/HIV patients who have a history of previous VL episodes relapse sooner and more often and are more likely to die than those presenting with their first episode of VL. The poorer prognosis of R as compared to P VL/HIV patients is accompanied by lower weight gain and recovery of WBC, RBC and PLT counts; and lower production of antigen specific IFNγ, lower CD4^+^ T cell counts and higher CD4 and CD8 PD1 iMFI. Low CD4^+^ T cell counts at end of treatment and high parasite load at time of diagnosis are both associated with higher risk of VL relapse in survival models, but neither of these associations remained significant when adjusted for previous VL history, which remained independently associated with relapse risk. This suggests that neither of these factors are the sole drivers of continued relapse in recurrent VL patients, but emphasises that a more complex immune dysfunction likely underlies the poor prognosis of recurrent VL.

Given the poor outcome predicted by VL/HIV coinfection and a history of relapse, specific measures should be tried to improve the long-term prognosis of these patients: an improvement of their ART treatment, such as inclusion of dolutegravir; more ART adherence counselling; a better follow-up of their nutritional status; and longer anti-leishmanial treatment. Additional molecular monitoring to inform the duration of anti-leishmanial treatment should be explored in future studies. Immune therapy, through PD1/PDL-1 blockade, that might improve the impaired production of antigen-specific IFNγ and/or through IFNγ administration (55) could result in more efficient parasite killing in these patients. Such interventions might contribute to the prevention of further relapse.

## AUTHOR CONTRIBUTIONS

All authors discussed the results and contributed to the final manuscript. Study conception and design: PK, YT, IM; Acquisition of data: YT, EA, TM, PK; Analysis and interpretation of data: YT, EA, RW, MK, ML, SUF, GPT, IM, JAC, PK; Drafting of manuscript: PK, YT, GPT, JAC, IM

## Data Availability

All data produced in the present work are contained in the manuscript

## ACKNOWLEDGMENTS

We are grateful to the staff of the Leishmaniasis Research and Treatment Centre for their support and DNDi for supporting the VL treatment service at the University of Gondar. We also would like to thank Prof. Charles Bangham for helpful discussion and critical reading of the manuscript; and Mandy Sanders and Siobhan Austin-Guest at the Wellcome Sanger Institute for supporting and co-ordinating sequencing work. YT is funded by a Wellcome Trust Training Fellowship in Public Health and Tropical Medicine (204797/Z/16/Z). This research was funded in part by the Wellcome Trust Grant [Grant number 206194, JAC]. For the purpose of Open Access, the authors have applied a CC BY public copyright licence to any Author Accepted Manuscript version arising from this submission. MK is funded by a Wellcome Trust Sir Henry Wellcome Fellowship (206508/Z/17/Z).

